# A meta-analysis on the role of pre-existing chronic disease in the cardiac complications of SARS-CoV-2 infection

**DOI:** 10.1101/2020.06.21.20136622

**Authors:** Jane E. Sinclair, Yanshan Zhu, Gang Xu, Wei Ma, Haiyan Shi, Kun-Long Ma, Chun-Feng Cao, Ling-Xi Kong, Ke-Qiang Wan, Juan Liao, Hai-Qiang Wang, Matt Arentz, Meredith Redd, Linda A. Gallo, Kirsty R. Short

## Abstract

**Background:** Severe acute respiratory syndrome coronavirus 2 (SARS-CoV-2) has been associated with multiple direct and indirect cardiovascular complications. It is not yet well defined whether the patient groups at increased risk of SARS-CoV-2 severe respiratory disease experience similarly heightened incidence of cardiovascular complications. We sought to analyse the role of host co-morbidities (chronic respiratory illnesses, cardiovascular disease (CVD), hypertension or diabetes mellitus (DM)) in the acute cardiovascular complications associated with SARS-CoV-2 infection.

**Methods:** We retrospectively investigated published and pre-printed, publicly released, de-identified, data made available between Dec 1, 2019, and October 14, 2020. Information was accessed from PubMed, Embase, medRxiv and SSRN. 363 full-text articles were reviewed and 283 excluded for lack of original research, irrelevance to outcome, inappropriate cohort, or describing case reports of <10 patients. Data were extracted from 25 studies and the remaining 55 contacted to request appropriate data, to which four responded with data contributions. A final of 29 studies were included. This systematic review was conducted based on Preferred Reporting Items for Systematic Reviews (PRISMA) and Meta-analyses of Observational Studies in Epidemiology (MOOSE) statements. Included studies were critically appraised using Newcastle Ottawa Quality Assessment Scale (NOS). Data were extracted independently by multiple observers. When *I* ^2^□<□50%, a fixed effects model was selected, while a random-effects model was employed when *I* ^2^□> 50%. Cardiovascular complications were measured as blood levels of cardiac biomarkers above the 99th percentile upper reference limit, new abnormalities in electrocardiography, and/or new abnormalities in echocardiography.

**Results:** Individual analyses of the majority of studies found that the median age was higher by approximately 10 years in patients with cardiovascular complications. Pooled analyses showed the development of SARS-CoV-2 cardiovascular complications was significantly increased in patients with chronic respiratory illness (Odds Ratio (OR) 1.67[1.48,1.88]), CVD (OR 3.37[2.57,4.43]), hypertension (OR 2.68[2.11,3.41]), DM (OR 1.60[1.31,1.95]) and male sex (OR 1.31[1.21,1.42]), findings that were mostly conserved during sub-analysis of studies stratified into four global geographic regions.

**Conclusions:** Age, chronic respiratory illness, CVD, hypertension, DM and male sex may represent prognostic factors for the development of both acute and chronic cardiovascular complications in COVID-19 disease, highlighting the need for a multidisciplinary approach to chronic disease patient management.

## INTRODUCTION

Severe acute respiratory syndrome coronavirus 2 (SARS-CoV-2) emerged at the end of 2019 and has since gone on to cause the ongoing coronavirus disease 2019 (COVID-19) pandemic, infecting >50 million people worldwide. SARS-CoV-2 causes a broad range of respiratory symptoms, ranging from subclinical disease to viral pneumonia [1]. However, it is increasingly apparent that SARS-CoV-2 also causes a variety of extra-respiratory complications [1]. Specifically, SARS-CoV-2 infection has been associated with acute myocardial injury, myocarditis, arrhythmias, venous thromboembolism and other cardiac complications [2–6]. The development of these complications is likely enhanced by experimental testing of antimalarial and antiviral drugs, which have known cardiovascular side effects [4–12]. Although prevalence estimates vary, data from China suggests that approximately 7% of patients infected with SARS-CoV-2 develop cardiac injury (defined as elevated high-sensitivity cardiac troponin I or new echocardiographic; electrocardiographic (ECG) abnormalities) [13]. This prevalence increases to 25% in those hospitalised with COVID-19 [14], and 22% in patients who required intensive care unit (ICU) admission [13]. However, at present, the role of pre-existing chronic disease in the cardiac complications of COVID-19 remains to be elucidated.

Pre-existing chronic respiratory disease, cardiovascular disease (CVD), hypertension and diabetes mellitus (DM) have each been associated with increased rates of hospitalization, ICU admission, and death from the respiratory complications of SARS-CoV-2 [13–27]. It is not yet well defined whether these patient groups are also at an increased risk of SARS-CoV-2 associated cardiac injury. Some small studies have shown that patients with pre-existing CVD are at a higher risk of myocarditis and/or myocardial infarction (MI) from SARS-CoV-2 [3, 28, 29]. A retrospective, single-centre case series of 187 patients with COVID-19 also found that pre-existing CVD increased the risk of cardiac injury (as defined by troponin elevation) (54.5% vs 13.2%) and mortality compared with patients without CVD [3]. Similarly, a meta-analysis of 28 observational studies found SARS-CoV-2 associated cardiac injury biomarkers to be related to a history of hypertension but not DM [30]. In contrast, Zhu and colleagues found that individuals with type 2 DM had a greater occurrence of acute heart injury (7.3% versus 3.0%) than individuals without DM [31].

There is a clear need to better understand the role of patient host factors in the extra-respiratory complications of SARS-CoV-2 infection. Here, we analysed the currently available literature on the role of pre-existing chronic respiratory illnesses, CVD, hypertension and DM in the prevalence of SARS-CoV-2 associated cardiac injury.

## METHODS

### Study selection

We retrospectively investigated published and pre-printed, publicly released and de-identified data made available between Dec 1, 2019, and October 14, 2020. Information was accessed from four databases (PubMed, Embase, medRxiv and SSRN) using the following search terms: (“covid-19”[All Fields]) OR (“SARS-CoV-2”[All Fields]) AND (“cardiac”[All Fields] AND “comorbid*”[All Fields]). A total of 1076 articles were retrieved after duplicates were removed (available in either English or Chinese), following which 713 records were removed after screening by title and abstract. 363 full-text articles were reviewed. Of these, 283 articles were excluded because i) they did not include original research, ii) they were not relevant to the outcome, iii) they described an inappropriate patient population and/or iv) they described case reports of <10 patients. Of the remaining 80 studies that were deemed eligible, due to their containing data on patient pre-existing disease and cardiac complications as an outcome, 25 contained the required data format and these were extracted. The authors of the remaining 55 studies were contacted to request appropriate data (as they presented only aggregate data that did not specify the cross-over between pre-existing chronic disease and outcome), to which four groups responded with data contributions. Thus, a final of 29 studies were included in the meta-analysis [3,4,29,32-57]. The selection process is shown in Figure 1.

**Figure 1.**
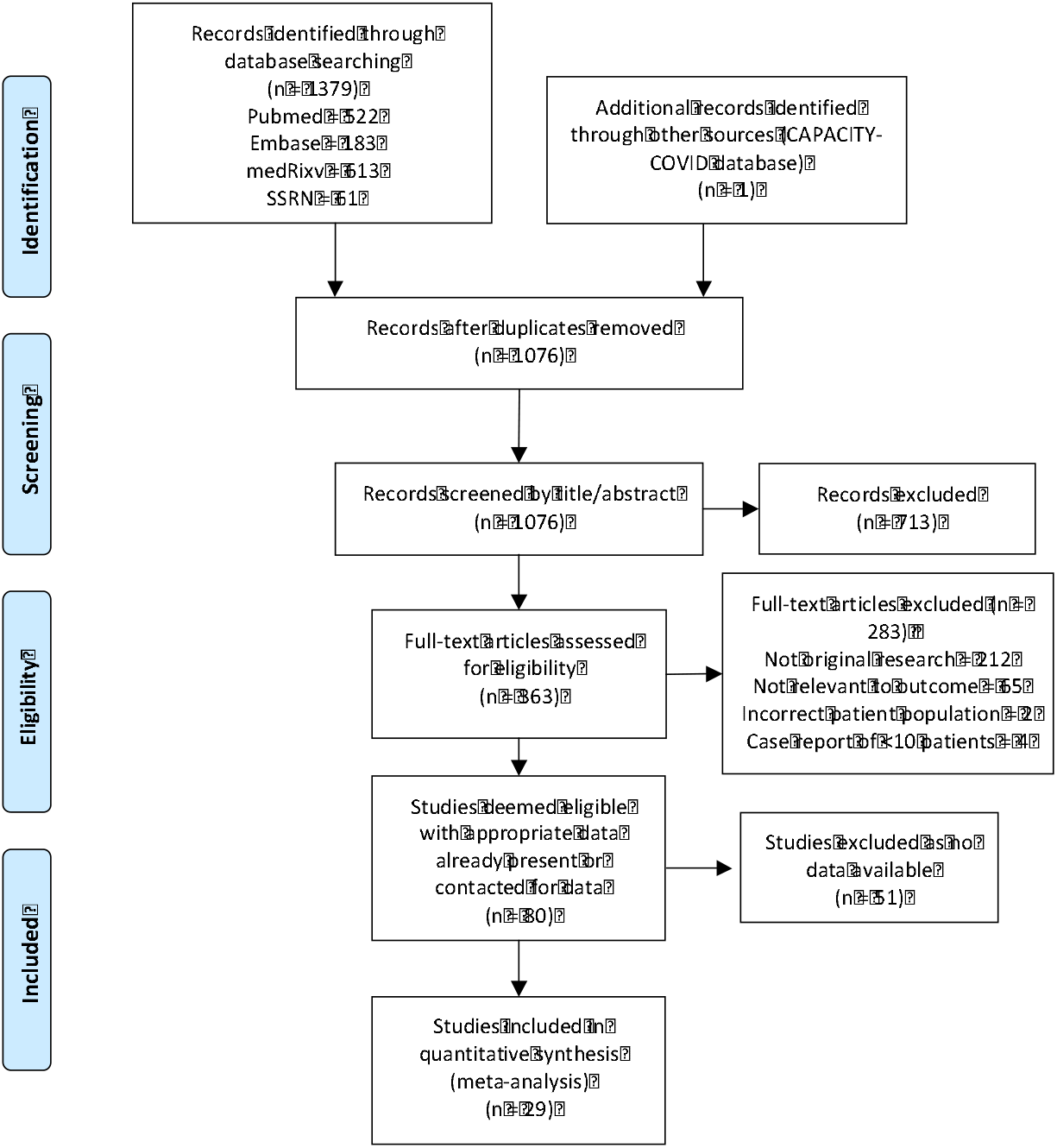
The Preferred Reporting Items for Systematic Reviews and Meta-analyses (PRISMA) flowchart of study selection.

### Data extraction and analysis

Extracted variables were aggregate data of median age, sex, underlying chronic disease (chronic respiratory illness, CVD, hypertension and DM) and the occurrence of cardiac complications as an outcome. Patients with co-morbidities were included in all applicable categories. For cases in which the data source reported more than one type of co-morbid CVD separately (such as prevalence of both chronic heart failure and coronary heart disease within their cohort), only the group with the largest number of cases was included to avoid patient double up and overrepresentation of co-morbid CVD. Analyses for each co-morbidity were performed independently, with each assuming one effect (the patient group in question) is considered as the main driver of effects. Median data for age were analysed individually using a Mann-Whitney U test. R version 4·0·0 software was used to analyse sex distribution as well as pre-existing chronic disease and cardiac outcomes through meta□analysis. Heterogeneity among studies was assessed by using the Q statistic with \**P*≤0.05 indicating the presence of significant heterogeneity and quantified by using the *I* ^2^ statistic. A fixed-effects model was used as when *I* ^2^□<□50% and random-effects model when *I* ^2^□>□50%.

### Definition of cardiac complications

Cardiac complications were defined as one or more of the following: blood levels of cardiac biomarkers (troponin I or creatine-kinase myocardial band) above the 99^th^ percentile upper reference limit [58–60], and/or abnormalities observed in clinical assessments of the patients’ heart function, including ECG [61–65] or echocardiography [62–66].

### Quality assessment and data synthesis

The included studies were critically appraised using Newcastle Ottawa Quality Assessment Scale (NOS) [67]. High-quality studies were defined as studies fulfilling NOS score of minimum 7. Data were synthesised from 29 different and high-quality studies [3,4,29,32–57]. This systematic review was conducted based on the Preferred Reporting Items for Systematic Review and Meta-Analysis (PRISMA) [68] and Meta-analysis of Observational Studies in Epidemiology (MOOSE) [69] statements.

## RESULTS

### SARS-CoV-2 patients with pre-existing chronic illnesses are at increased risk of acute cardiac complications

We identified 1076 articles that described pre-existing chronic disease and cardiac complications of SARS-CoV-2, rejected 996 articles due to a lack of sufficient and/or appropriate data and derived a total of 80 articles. 29 of these studies were subsequently used in a meta-analysis (Figure 1), all of which were high quality (Table 1). Baseline characteristics of the patient populations as well as other relevant data are listed in Table 2. The definition of cardiac complications varied between studies, ranging from acute cardiac injury to fulminant myocarditis. 20 studies measured cardiac outcomes on the day of admission [3,29,32,34,35,37–41,43–48,50-52,57], and one at the time of patient death [33], with the remaining eight studies not specifying the time of assessment [4,36,42,49,53–55]. The studies were conducted in 10 countries in four geographical locations (U.S.A. [32,33,41,48,49], the Middle East [34,50,52], Europe [4,35–40,44,45,47], and the People’s Republic of China [3,29,42,43,46,51,53–57]) with a total of 3553 COVID-19 patients developing cardiac complications and 13,225 COVID-19 patients without cardiac complications. Patients who presented with SARS-CoV-2 associated cardiac complications were, on average, 10 years older than in those who did not (Table 2).

**Table 1.** Quality assessment of the included studies using the Newcastle Ottawa Scale.

| Study | Selection | Comparability | Outcome | Total |
| --- | --- | --- | --- | --- |
| Abrams et al., 2020<sup>32</sup> | **** | -- | *** | 7 |
| Arentz et al., 2020<sup>33</sup> | **** | -- | *** | 7 |
| Barman et al., 2020<sup>34</sup> | **** | -- | *** | 7 |
| Bertini et al., 2020<sup>35</sup> | **** | -- | *** | 7 |
| CAPACITY COVID, 2020<sup>36</sup> | **** | -- | *** | 7 |
| Duerr et al., 2020<sup>37</sup> | **** | -- | *** | 7 |
| Ferrante et al., 2020<sup>38</sup> | **** | -- | *** | 7 |
| Ghio et al., 2020<sup>39</sup> | **** | -- | *** | 7 |
| Guo et al., 2020<sup>3</sup> | **** | -- | *** | 7 |
| Lairez et al., 2020<sup>40</sup> | **** | -- | *** | 7 |
| Lala et al., 2020<sup>41</sup> | **** | -- | *** | 7 |
| Li et al., 2020<sup>42</sup> | **** | -- | *** | 7 |
| Liu et al., 2020<sup>43</sup> | **** | -- | *** | 7 |
| Lombardi et al., 2020<sup>44</sup> | **** | -- | *** | 7 |
| Lorente-Ros et al., 2020<sup>45</sup> | **** | -- | *** | 7 |
| Ma et al., 2020<sup>46</sup> | **** | -- | *** | 7 |
| Pagnesi et al., 2020<sup>47</sup> | **** | -- | *** | 7 |
| Raad et al., 2020<sup>48</sup> | **** | -- | *** | 7 |
| Sala et al., 2020<sup>4</sup> | **** | -- | *** | 7 |
| Shah et al., 2020<sup>49</sup> | **** | -- | *** | 7 |
| Shahrokh et al., 2020<sup>50</sup> | **** | -- | *** | 7 |
| Shi et al., 2020<sup>29</sup> | **** | -- | *** | 7 |
| Song et al., 2020<sup>51</sup> | **** | -- | *** | 7 |
| Szekely et al., 2020<sup>52</sup> | **** | -- | *** | 7 |
| Toraih et al., 2020<sup>53</sup> | **** | -- | *** | 7 |
| Wei et al., 2020<sup>54</sup> | **** | -- | *** | 7 |
| Xie et al., 2020<sup>55</sup> | **** | -- | *** | 7 |
| Xu G et al., 2020<sup>56</sup> | **** | -- | *** | 7 |
| Xu H et al., 2020<sup>57</sup> | **** | -- | *** | 7 |

**Table 2.**
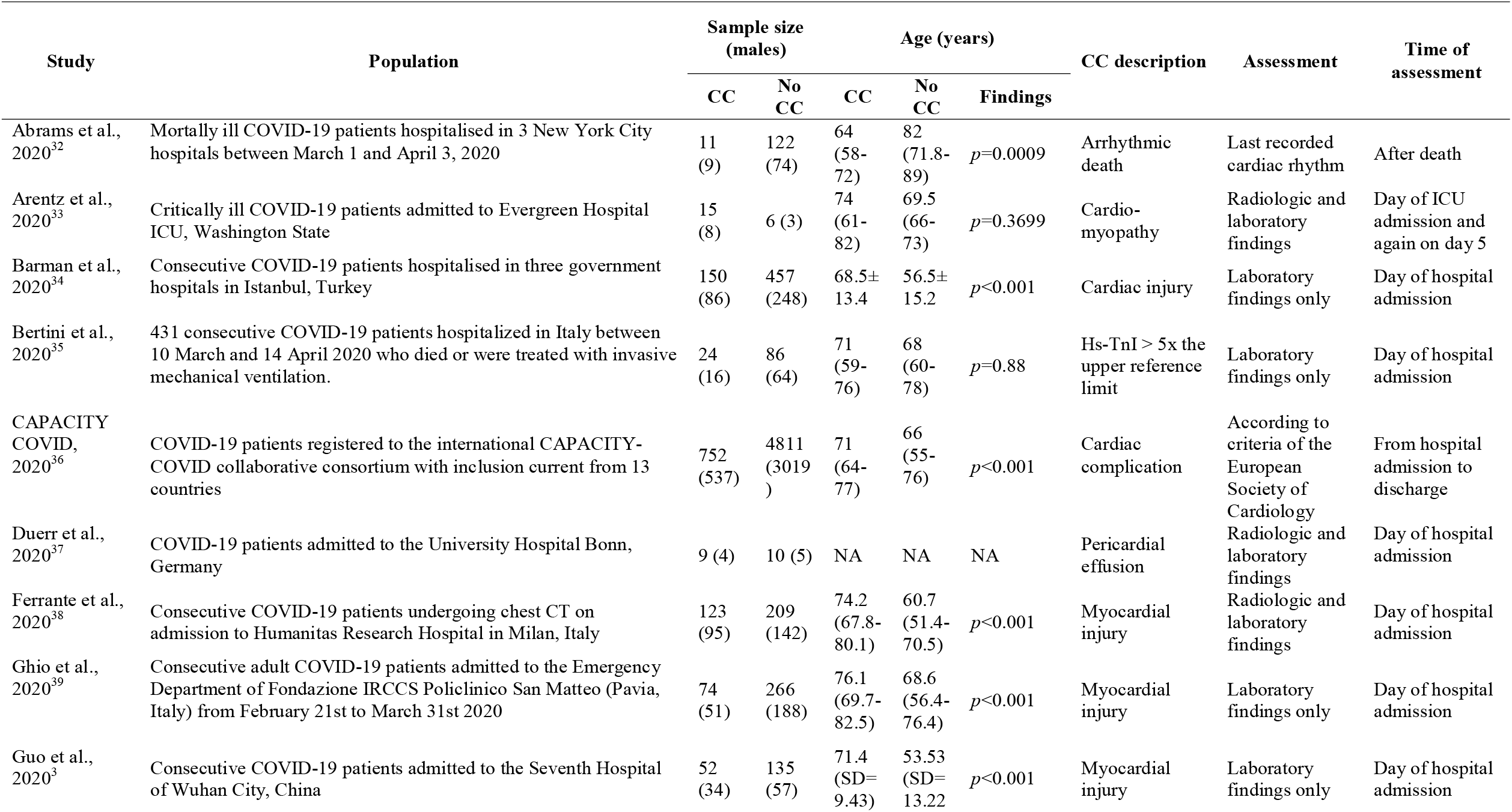
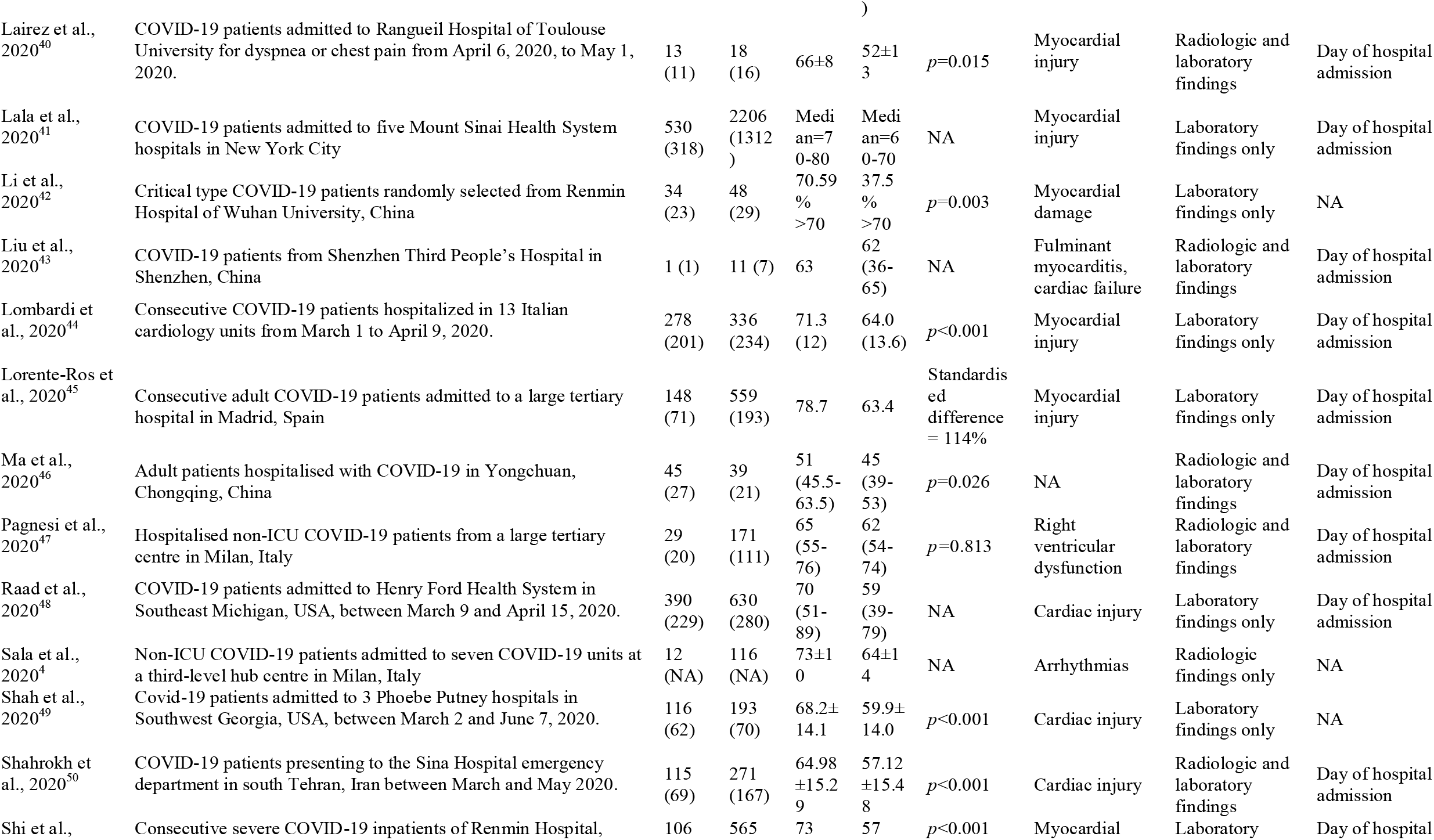
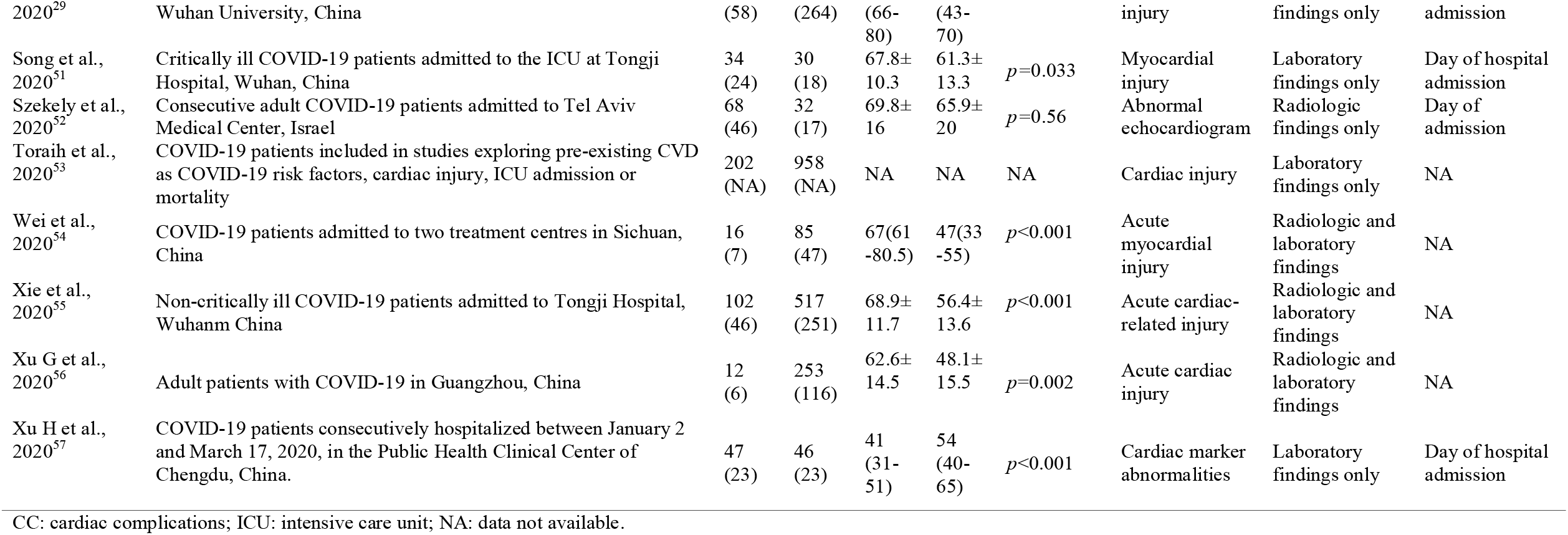
Summary of baseline characteristics of the included studies.

In a pooled analysis, chronic respiratory illness (odds ratio (OR): 1.67, 95% confidence interval (CI) [1.48,1.88]) (Fig. 2), CVD (OR: 3.37, 95% CI [2.57,4.43]) (Fig. 3), hypertension (OR: 2.68, 95% CI [2.11,3.41]) (Fig. 4), DM (OR: 1.60, 95% CI [1.31,1.95]) (Fig. 5), and male sex (OR: 1.31, 95% CI [1.21,1.42]) (Fig. 6), were each associated with a higher incidence of SARS-CoV-2 associated cardiac complications.

**Figure 2:**
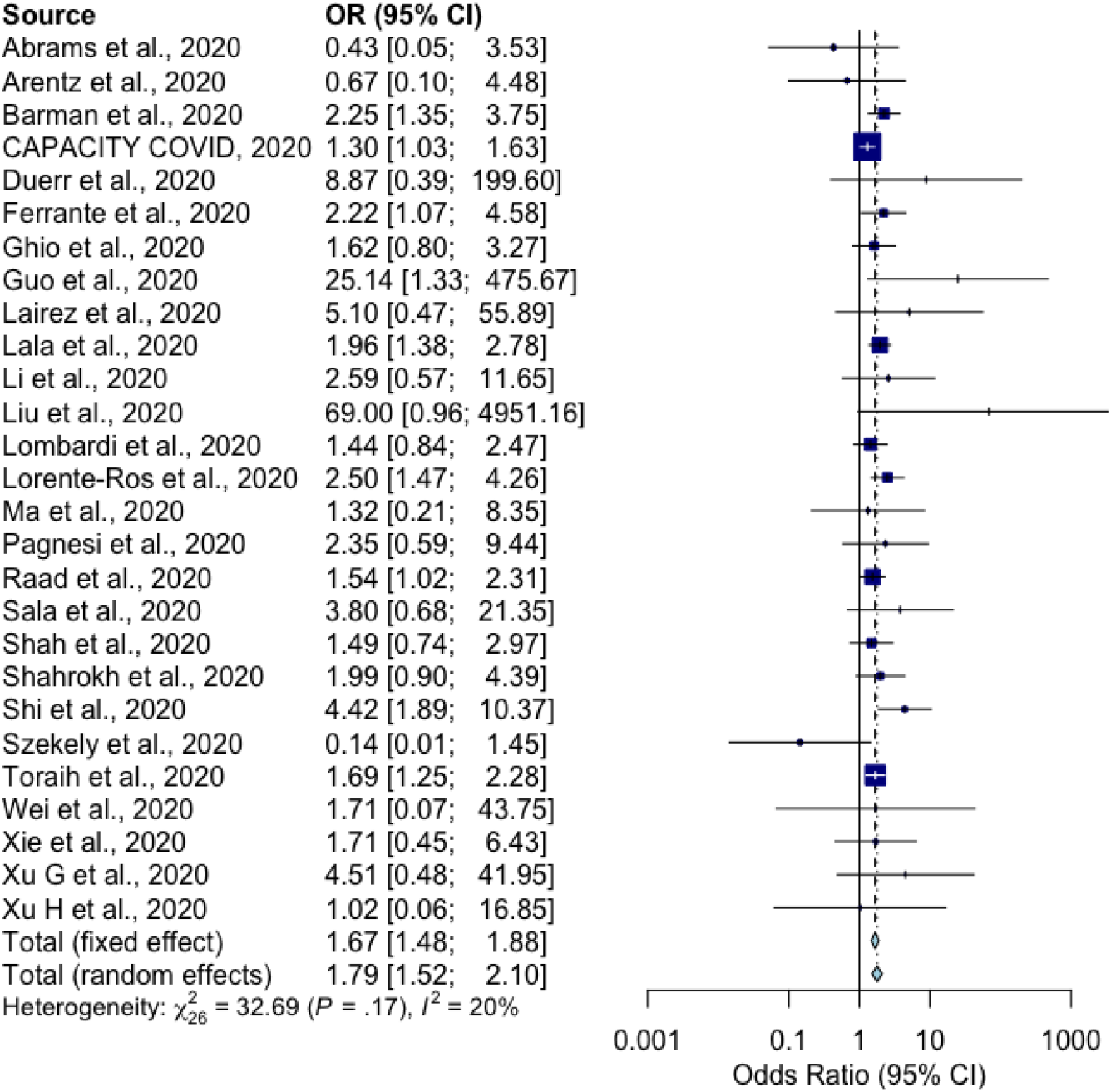
Forest plot of chronic respiratory illness as a risk factor for cardiac complications in COVID-19 patients. Fixed effect models apply when *I*^2^<50%.

**Figure 3:**
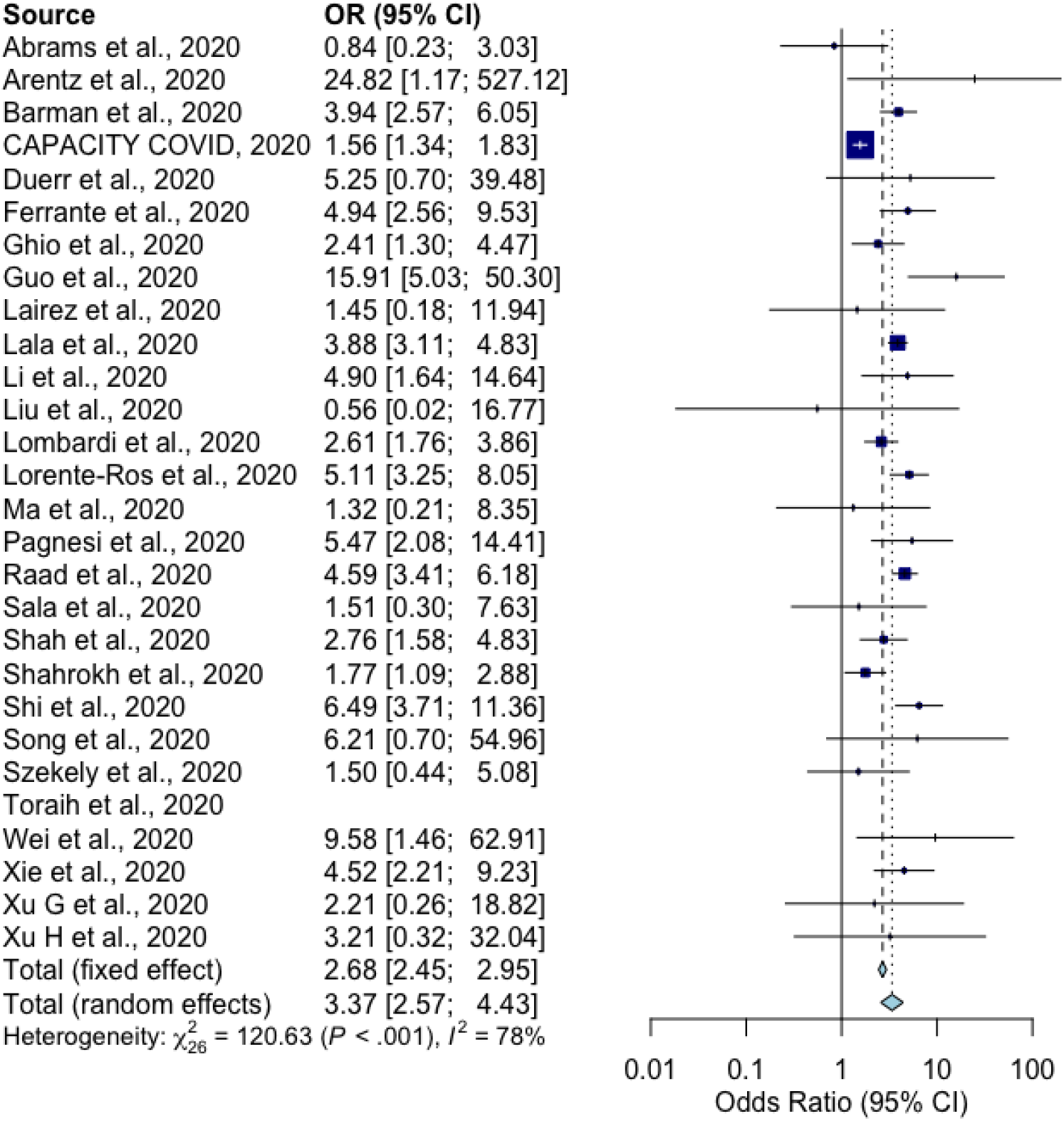
Forest plot of pre-existing cardiovascular disease as a risk factor for cardiac complications in COVID-19 patients. Random effect models apply when *I*^2^>50%.

**Figure 4:**
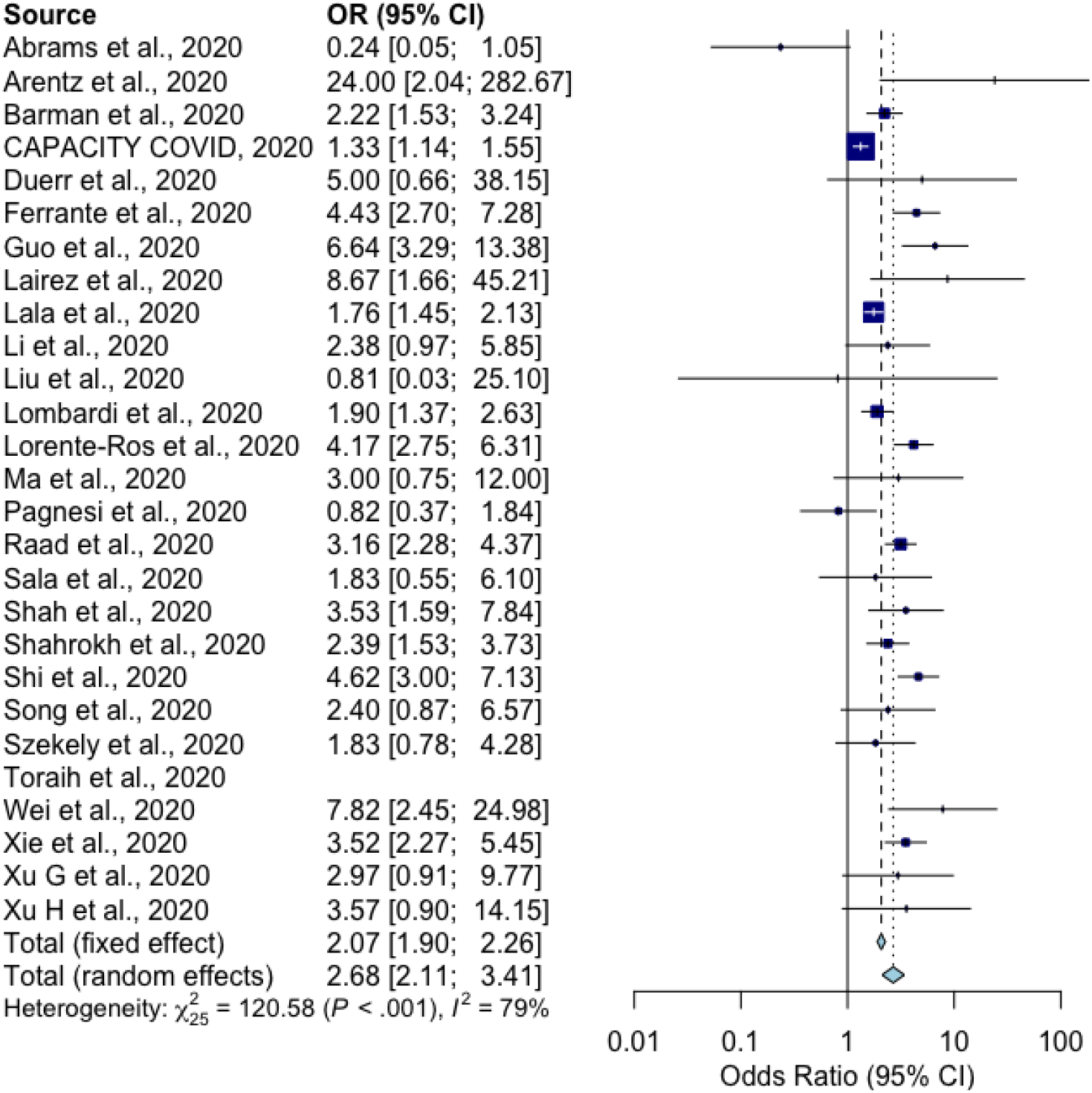
Forest plot of hypertension as a risk factor for cardiac complications in COVID-19 patients. Random effect models apply when *I*^2^>50%.

**Figure 5:**
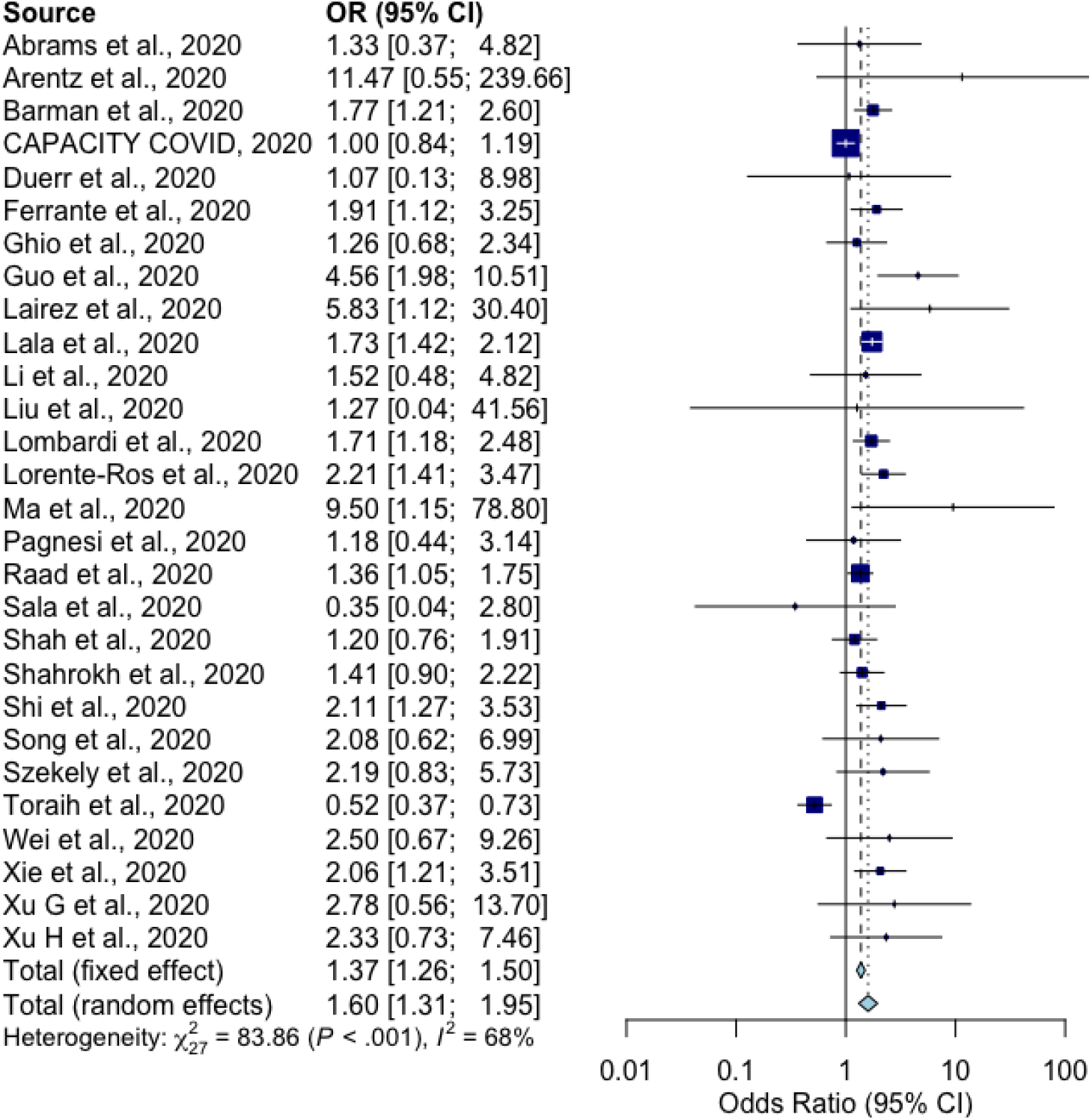
Forest plot of diabetes mellitus as a risk factor for cardiac complications in COVID-19 patients. Random effect models apply when *I*^2^>50%.

**Figure 6:**
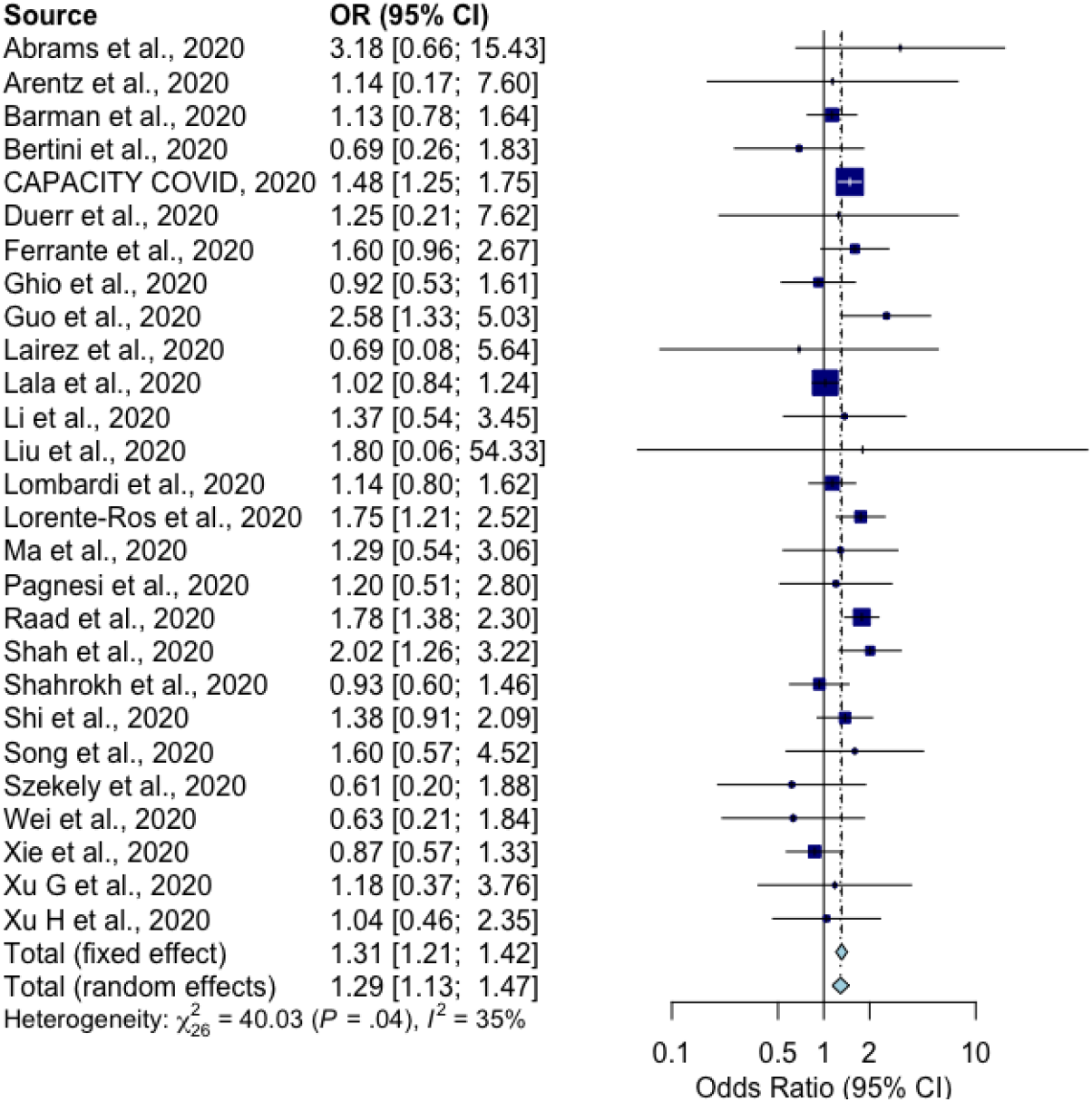
Forest plot of male sex as a risk factor for cardiac complications in COVID-19 patients. Fixed effect models apply when *I*^2^<50%.

As large heterogeneity was detected amongst the total combined study cohort, the cohort was subdivided based on general global region in an attempt to account for this. Cohorts were divided into studies with cohorts primarily situated in the U.S.A [32,33,41,48,49], the Middle East [34,50,52], Europe [4,35–40,44,45,47], and China [3,29,42,43,46,51,53–57].

Regionally-pooled sub-analysis of chronic respiratory illness found it to be associated with a higher incidence of SARS-CoV-2 associated cardiac complications in cohorts from the U.S.A. (OR: 1.64, 95% CI [1.28,2.09]) (Fig. 7, A), Europe (OR: 1.53, 95% CI [1.28,1.83]) (Fig. 7, C) and China (OR: 1.94, 95% CI [1.49,2.52]) (Fig. 7, D), but not the Middle East (Fig. 7, B). Regional subdivision accounted for the high heterogeneity amongst all but the Middle Eastern cohort.

**Figure 7:**
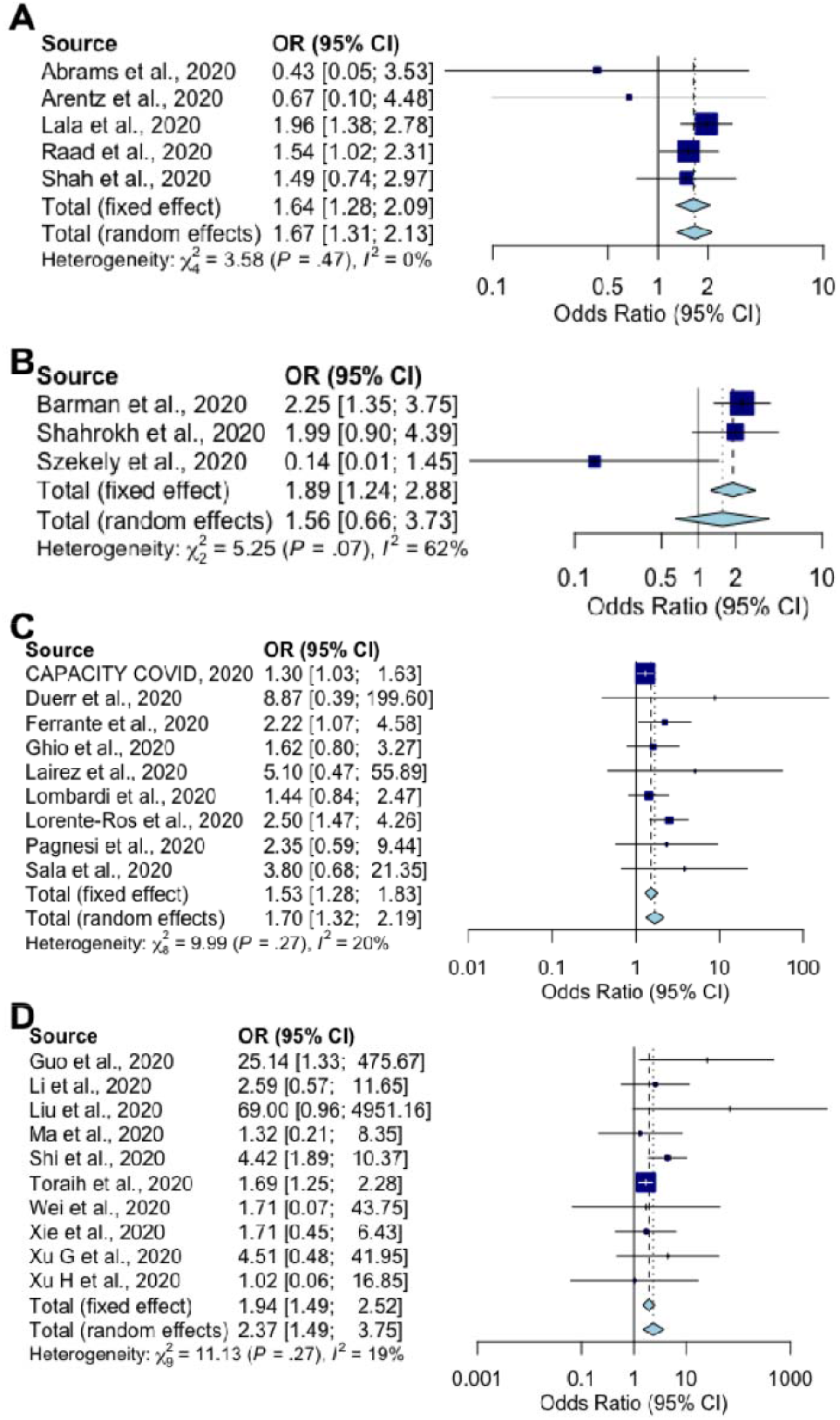
Forest plots of chronic respiratory as a risk factor for cardiac complications in COVID-19 patients divided by geographical region. A | Studies based primarily in the United States of America. B | Studies based primarily around the Middle East. C | Studies based primarily in Europe. D | Studies based primarily in China. Fixed effect models apply when *I*^2^<50% and random effects models apply when *I*^2^>50%.

Regionally-pooled sub-analysis of pre-existing CVD found its association with a higher incidence of SARS-CoV-2 associated cardiac complications to be conserved across all cohorts from the U.S.A. (OR: 3.55, 95% CI [2.51,5.04]) (Fig. 8, A), the Middle East (OR: 2.40, 95% CI [1.25,4.62]) (Fig. 8, B), Europe (OR: 2.99, 95% CI [1.90,4.69]) (Fig. 8, C), and China (OR: 5.52, 95% CI [3.88,7.85]) (Fig. 8, D), despite regional subdivision failing to account for high heterogeneity amongst the former three regions.

**Figure 8:**
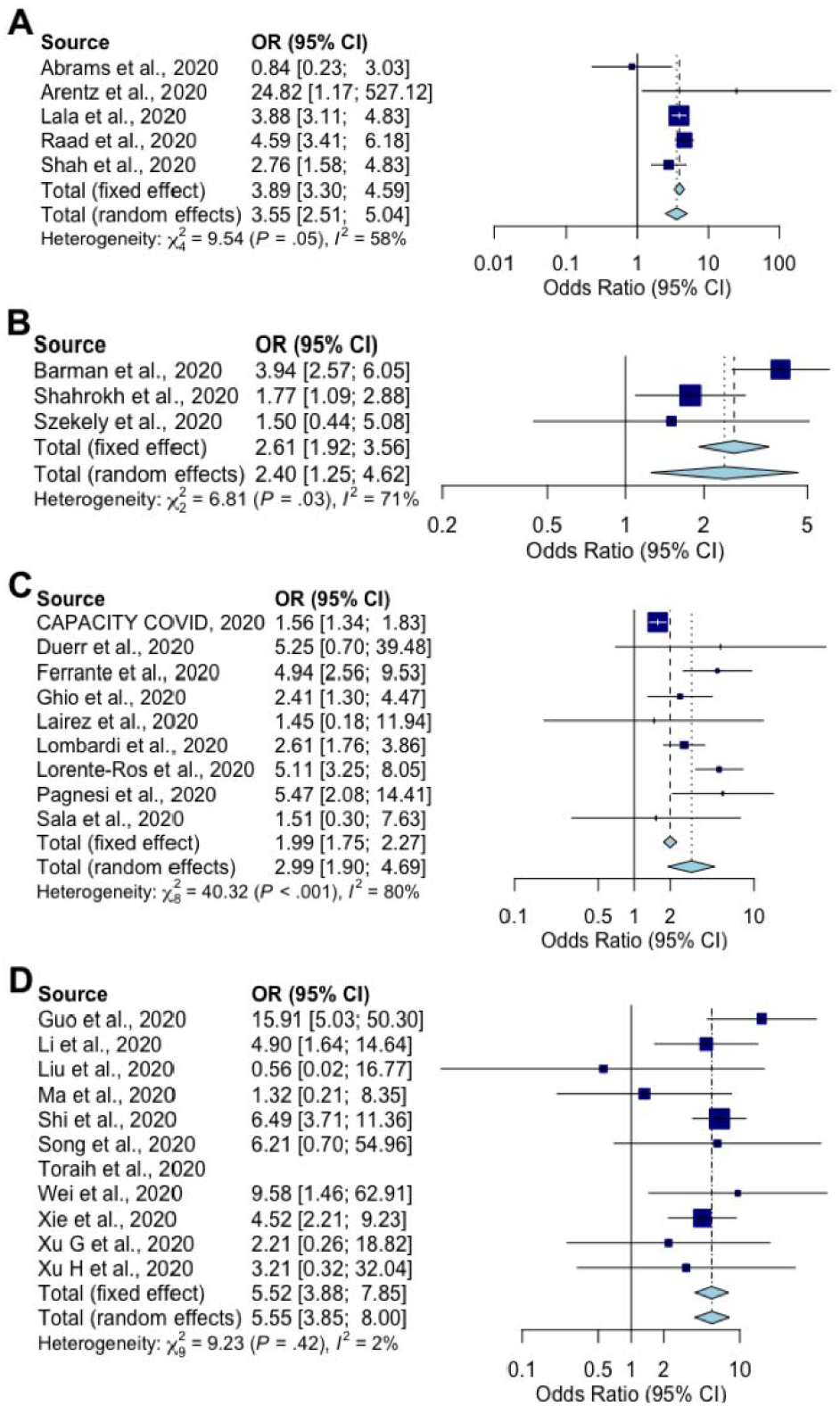
Forest plots of pre-existing cardiovascular disease as a risk factor for cardiac complications in COVID-19 patients divided by geographical region. A | Studies based primarily in the United States of America. B | Studies based primarily around the Middle East. C | Studies based primarily in Europe. D | Studies based primarily in China. Fixed effect models apply when *I*^2^<50% and random effects models apply when *I*^2^>50%.

Regionally-pooled sub-analysis of hypertension found its association with a higher incidence of SARS-CoV-2 associated cardiac complications to be similarly conserved across all cohorts from the U.S.A. (OR: 2.23, 95% CI [1.22,4.08]) (Fig. 9, A), the Middle East (OR: 2.24, 95% CI [1.70,2.94]) (Fig. 9, B), Europe (OR: 2.38, 95% CI [1.46,3.88]) (Fig. 9, C), and China (OR: 3.94, 95% CI [3.10,5.01]) (Fig. 9, D), although regional subdivision could not account for high heterogeneity amongst the American and European cohorts.

**Figure 9:**
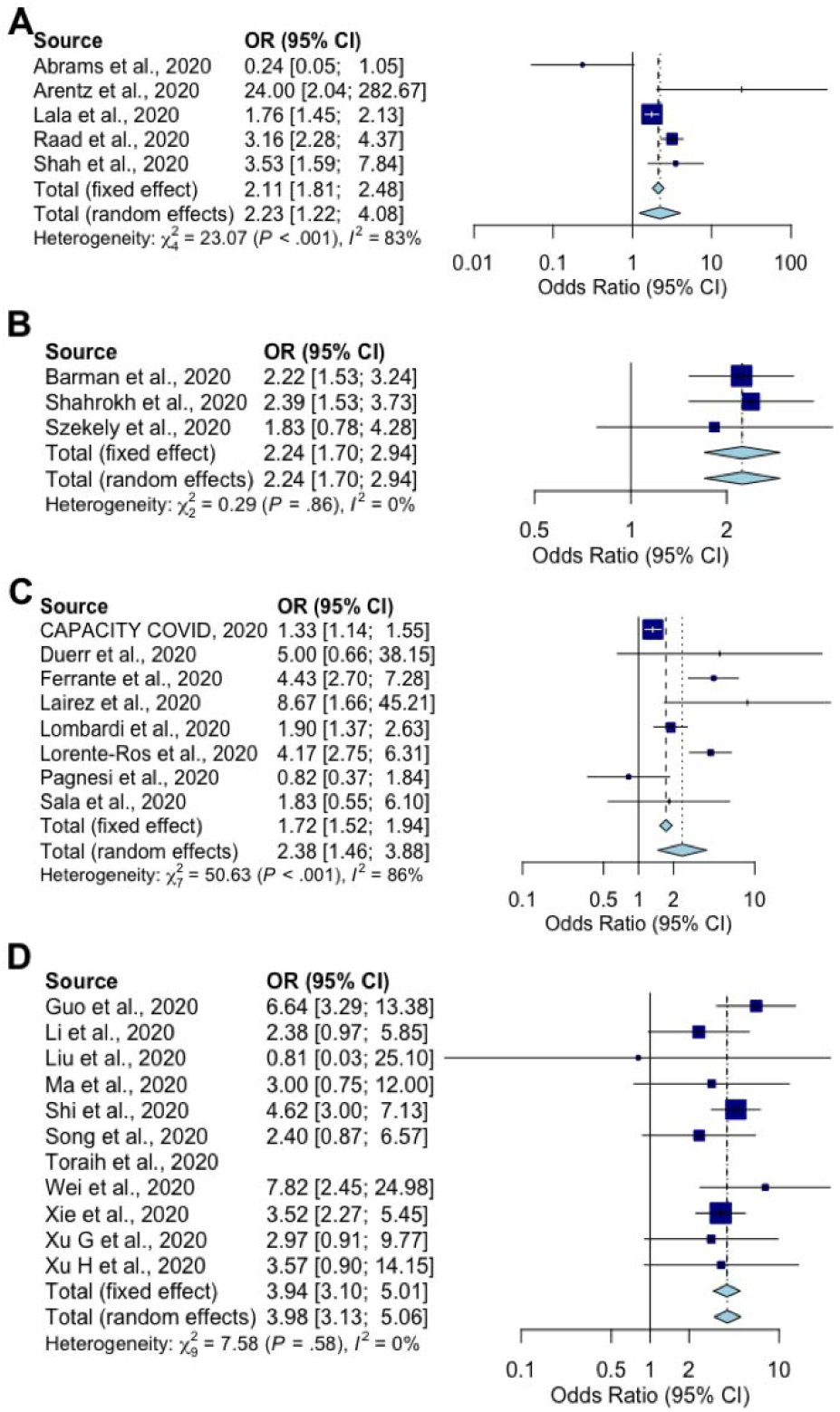
Forest plots of hypertension as a risk factor for cardiac complications in COVID-19 patients divided by geographical region. A | Studies based primarily in the United States of America. B | Studies based primarily around the Middle East. C | Studies based primarily in Europe. D | Studies based primarily in China. Fixed effect models apply when *I*^2^<50% and random effects models apply when *I*^2^>50%.

The association between DM and a higher incidence of SARS-CoV-2 associated cardiac complications was also conserved across all cohorts from the U.S.A. (OR: 1.54, 95% CI [1.32,1.78]) (Fig. 10, A), the Middle East (OR: 1.65, 95% CI [1.25,2.19]) (Fig. 10, B), Europe (OR: 1.50, 95% CI [1.07,2.09]) (Fig. 10, C), and China (OR: 2.04, 95% CI [1.13,3.70]) (Fig. 10, D), despite regional subdivision not accounting for high heterogeneity amongst the European and Chinese cohorts.

**Figure 10:**
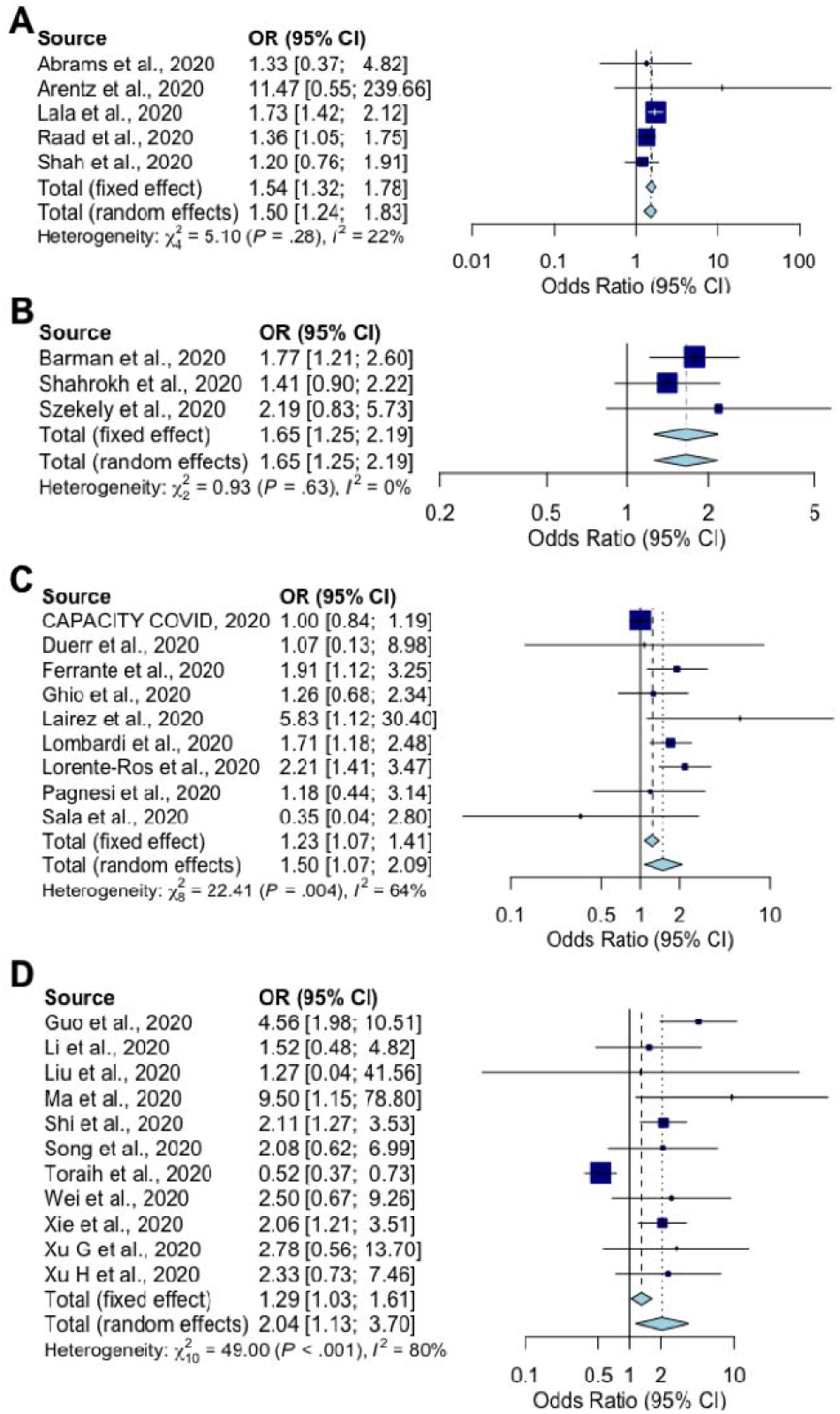
Forest plots of diabetes mellitus as a risk factor for cardiac complications in COVID-19 patients divided by geographical region. A | Studies based primarily in the United States of America. B | Studies based primarily around the Middle East. C | Studies based primarily in Europe. D | Studies based primarily in China. Fixed effect models apply when *I*^2^<50% and random effects models apply when *I*^2^>50%.

Finally, regionally-pooled sub-analysis of male sex found it to be associated with a higher incidence of SARS-CoV-2 associated cardiac complications in cohorts from the U.S.A. (OR: 1.55, 95% CI [1.03,2.33]) (Fig. 11, A), and Europe (OR: 1.40, 95% CI [1.23,1.59]) (Fig. 11, C), but not the Middle East (Fig. 11, B) or China (Fig. 11, D). Regional subdivision accounted for the high heterogeneity amongst all but the American cohort.

**Figure 11:**
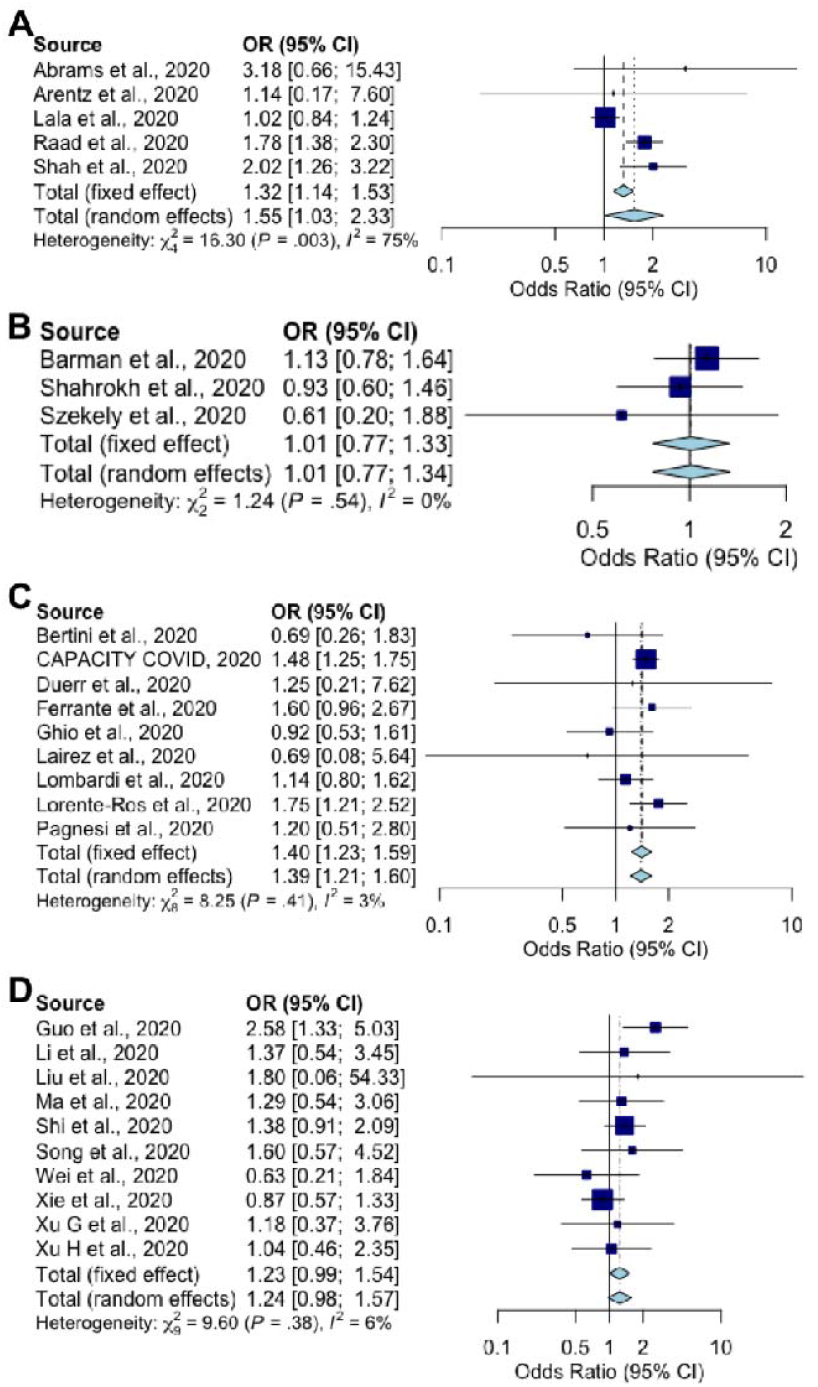
Forest plots of male sex as a risk factor for cardiac complications in COVID-19 patients divided by geographical region. A | Studies based primarily in the United States of America. B | Studies based primarily around the Middle East. C | Studies based primarily in Europe. D | Studies based primarily in China. Fixed effect models apply when *I*^2^<50% and random effects models apply when *I*^2^>50%.

## DISCUSSION

Our meta-analysis shows that older age, male sex, and pre-existing chronic respiratory disease, CVD, hypertension and DM are all risk factors for the development of cardiac complications associated with SARS-CoV-2 infection. At present the reasons for each of these association are unclear; each is a respiratory risk factor and thus might cause an indirect cardiac effect, however it could equally be the case that they exert their own direct catalytic effect on cardiac complications.

Our analysis identified a significant association between pre-existing CVD and the risk of SARS-CoV-2 associated cardiac complications, which was conserved across global geographical regions. This is consistent with previous observations that 30% of patients with SARS-CoV-2 cardiac injury had a history of coronary heart disease [3,29]. The reason for this association remains unclear. These data may be the result of pre-existing CVD leading to an altered immune response, excessive cytokine release [5], endothelial dysfunction, increased procoagulant blood activity, rupturing of coronary atherosclerotic plaques [73], and the formation of an occlusive thrombi [74].However, further investigations are required to validate this hypothesis.

The meta-analysis performed herein also found that chronic respiratory illness significantly increased the incidence of SARS-CoV-2 associated cardiac complications. It is possible that these data reflect the established indirect association between severe respiratory disease and subsequent acute cardiovascular event Indeed, COVID-19 patients with pneumonia and acute respiratory distress syndrome (ARDS) display acute right ventricular hypertrophy despite an absence of pulmonary embolism or severe pulmonary hypertension [87]. However, chronic lung disease has not been consistently associated with SARS-CoV-2 ARDS and hypoxemia [88], and thus this is unlikely to be the sole explanation for the increased risk of cardiac complications in patients with chronic respiratory illness in the present analysis. SARS-CoV-2 associated cardiac complications

The association between underlying DM and cardiac complications from SARS-CoV-2 may be attributable to a multitude of other effects induced by abnormal circulating substrates, including hyperglycaemia, and/or the co-existence and complex interactions with other, even subclinical, conditions in this patient group [91]. In response to changes in substrate availability, for example, the diabetic heart becomes increasingly dependent on fatty acids as a fuel source, which decreases work efficiency, and dysregulates responses to hypoxia [92]. These metabolic changes may predispose individuals with DM to an increased risk of ischemic cardiac injury upon SARS-CoV-2 infection. The contribution of underlying DM to the risk of SARS-CoV-2 associated cardiac complications, however, depends largely on the duration of pre-existing disease [93].

There are several limitations in our review. Firstly, the limited number of studies that met the pre-defined selection criteria disallowed correction for other variables that may confound these results. For example, it is possible that our observations were confounded by the strong effect that age has on cardiovascular health. Thus, it is not possible to conclude if people with these pre-existing conditions are just more susceptible to severe COVID-19 and thus the heart complications associated with this increased severity, or if there is indeed a cardiac-specific effect occurring. Similarly, we cannot exclude the possibility that the associations observed herein are due, at least in part, to treatments for severe COVID-19 that may damage the heart. Indeed, the experimental therapeutic hydroxychloroquine has been associated with QT interval prolongation, cardiomyopathy and heart failure [8–10]. This is likewise the case for treatments for underlying conditions, such as ACE inhibitors and ARBs which, while not known to damage the heart, may interfere with how the immune system targets the virus [78]. Information regarding which, if any, therapeutics used in these cohorts of patients was not consistently available. We anticipate that as more data on the cardiac complications become available it will be possible for future analyses to correct for these and other confounders; however, it would also be worthwhile testing the effects of each these pre-existing chronic conditions in one of the recently developed murine models of SARS-CoV-2.

Despite the above limitations, these data have important implications for patient care. Patients with one or more underlying medical conditions should be closely monitored for both severe respiratory and extra-respiratory COVID-19 complications. Furthermore, the exploratory use of medications that are independently associated with cardiac complications should be considered with caution in these patient groups and perhaps bypassed in favour of lower-risk treatment options. There is thus a clear need for a multidisciplinary, collaborative team to manage the complexity of COVID-19 illness observed in patients with one or more underlying chronic disease.

## RESOURCE AVAILABILITY

### Lead contact

Further information and requests for resources should be directed to and will be fulfilled by the Lead Contact, Dr. Kirsty Short.

### Materials availability

This study did not generate new unique reagents.

### Data and code availability

All data relevant to this study has been included in the manuscript. Data contributed by Xu et al., 2020^31^, Ma et al., 2020^32^, and Arentz et al., 2020^35^, may be obtained by contacting authors of each respective study. Data retrieved from remaining studies are published and publicly available. R version 4·0·0 software was used to analyse sex distribution as well as pre-existing chronic disease and cardiac outcomes through meta□analysis. The datasets/code supporting the current study have not been deposited in a public repository but are available from the corresponding author on request.

## Data Availability

All data relevant to this study has been included in the manuscript. Data contributed by Xu et al., Ma et al., and Arentz et al., may be obtained by contacting authors of each respective study. Data retrieved from remaining studies are published and publicly available.

## DECLARATIONS

## Acknowledgements

We want express our gratitude to the CAPACITY-COVID collaborative consortium for their data contribution:

Admiraal de Ruyter Hospital, Goes, the Netherlands: van Kesteren HAM

Albert Schweitzer Hospital, Dordrecht, the Netherlands: van de Watering DJ Amphia Hospital, Breda, the Netherlands: Schaap J, Alings AMW

Amstelland Hospital, Amstelveen, the Netherlands: Asselbergs-Koning AMH

Amsterdam University Medical Center, Amsterdam, the Netherlands: Pinto Y, Tjong FVY Antonius Hospital, Sneek, the Netherlands: de Vries JK

Antwerp University Hospital, Antwerp, Belgium: van Craenenbroeck EM, Kwakkel – van Erp HM, van Ierssel S AZ Maria Middelares, Gent, Belgium: de Sutter J

Barts Health NHS Trust, London, United Kingdom: Saxena M

Beatrix Hospital, Gorinchem, the Netherlands: Westendorp PHM Bernhoven Hospital, Uden, the Netherlands: van de Wal RMA

Bravis Hospital, Roosendaal, the Netherlands: Dorman HGR, van Boxem AJM Catharina Hospital, Eindhoven, the Netherlands: Tio RA

CHU UCL Namur – Site Godinne, Yvoir, Belgium: Gabriel LG Deventer Hospital, Deventer, the Netherlands: Badings EA

Diakonessenhuis, Utrecht, the Netherlands: van Ofwegen-Hanekamp CEE Dijklander Hospital, Hoorn, the Netherlands: Wierda E

Dutch Network for Cardiovascular Research (WCN): Schut A

Elizabeth-TweeSteden Hospital, Tilburg, the Netherlands: Monraats PS E.M.M.S Hospital, Nazareth, Israel: Hellou EH

Erasmus University Medical Center, Rotterdam, the Netherlands: den Uil CA, Jewbali LS

Fransiscus Gasthuis, Rotterdam, the Netherlands: Nierop PR

Fransiscus Vlietland, Schiedam, the Netherlands: van der Linden MMJM

Gelre Hospitals, Apeldoorn, the Netherlands: Groenemeijer BE

Gelre Hospitals, Zutphen, the Netherlands: Al-Windy NYY

Groene Hart Hospital, Gouda, the Netherlands: van Hessen MWJ

Haaglanden Medical Center, den Haag, the Netherlands: van der Heijden DJ

Hospital do Espirito Santo, Évora, Portugal: Ribeiro MIA

Hospital Group Twente, Almelo, the Netherlands: Linssen GCM

Hospital Prof. Doutor Fernando Fonesca, Amadora, Portugal: Ferreira JB

Imperial College Healthcare NHS Trust, London, United Kingdom: Prasad S

I.M. Sechenov First Moscow State Medical University, Moscow, Russia: Kopylov Ph. Yu., Blagova OV

Ikazia Hospital, Rotterdam, the Netherlands: Emans ME

Incliva Research Institute, University of Valencia, Valencia, Spain: Redón J, Forner MJ

Isala, Zwolle, the Netherlands: Hermanides RS

Jeroen Bosch Hospital, ‘s-Hertogenbosch, the Netherlands: Haerkens-Arends HE

Jessa Hospital, Hasselt, the Netherlands: Timmermans P Jr, Messiaen P

Julius Center for Health Sciences and Primary Care, University Medical Center Utrecht, The Netherlands: van Smeden M

King Fahd Hospital of the University, Khobar, Saudi Arabia: Al-Ali AK, Al-Muhanna FA, Al-Rubaish AM, Almubarak YA, Alnafie AN, Alshahrani M, Alshehri A

LangeLand Hospital, Zoetermeer, the Netherlands: van der Meer P

Leeds Teaching Hospitals NHS Trust, Leeds, United Kingdom: Kearney M

Leeuwarden Medical Center, Leeuwarden, the Netherlands: van den Brink FS

Leiden University Medical Center, Leiden, the Netherlands: Siebelink HJ

Maasstad Hospital, Rotterdam, the Netherlands: Smits PC

Maastricht University Medical Center, Maastricht, the Netherlands: Heymans SRB

Manchester University NHS Foundation Trust, Manchester, United Kingdom: Buch M

Martini Hospital, Groningen, the Netherlands: Tieleman RG, Reidinga AC

Meander Medical Center, Amersfoort, the Netherlands: Mosterd A

Medisch Spectrum Twente, Enschede, the Netherlands: Meijs MFL, Delsing CE, van Veen HPAA

Netherlands Heart Institute, Utrecht, the Netherlands: Hermans-van Ast W, Iperen EPA

Northumbria Healthcare NHS Foundation Trust, Cramlington, United Kingdom: Aujayeb A

One Day Surgery Hospital, Cairo, Egypt: Salah R

Queen Elizabeth University Hospital, Glasgow, United Kingdom: Touyz R

Rijnstate Hospital, Arnhem, the Netherlands: Seelig J

Rode Kruis Hospital, Beverwijk, the Netherlands: Westendorp ICD, Veldhuis LI

Royal Brompton and Harefield NHS Foundation Trust, London, United Kingdom: Prasad S

Royal Devon and Exeter NHS Foundation Trust, Exeter, United Kingdom: Shore AC

Royal Free London NHS Foundation Trust, London, United Kingdom: Captur G

Salford Royal NHS Foundation Trust, Salford, United Kingdom: Dark P

San Luigi Gonzaga University Hospital, Orbassano, Turin, Italy: Montagna LM, Bianco MB, Mazzilli SGM

Saxenburgh Medical Center, Hardenberg, the Netherlands: Kaplan R, Drost JT

Slingeland Hospital, Doetinchem, the Netherlands: Schellings DAAM

Southern Health and Social Care Trust, Portadown, United Kingdom: Moriarty A

Spaarne Gasthuis, Haarlem, the Netherlands: Kuijper AFM

SSR Val Rosay, Saint-Didier-au-Mont-d’Or, France: Charlotte NC St. Antonius Hospital, Nieuwegein, the Netherlands: ten Berg JM

St. Jansdal, Harderwijk, the Netherlands: van der Zee PM

Tehran Heart Center, Tehran, Iran: Shafiee A, Hedayat B, Poorhosseini H, Saneei E

The Newcastle upon Tyne Hospitals NHS Foundation Trust, Newcastle-upon-Tyne, United Kingdom: Zaman A

Treant Zorggroep, Hoogeveen, the Netherlands: Anthonio RL

University College London Hospitals NHS Foundation Trust, London, United Kingdom: Williams B, Patel R, Bell R

University Hospitals Bristol NHS Foundation Trust, Bristol, United Kingdom: Caputo M

University Hospital Brussels, Brussels, Belgium: Weytjens C

University Hospital Complex of Granada, Granada, Spain: Macías-Ruiz R University Hospital of Geneva, Geneva, Switzerland: Tessitore E

University Hospitals of Leicester NHS Trust, Leicester, United Kingdom: McCann GP

University Medical Center Groningen, Groningen, the Netherlands: van Gilst WH

University Medical Center Utrecht, Utrecht, the Netherlands: Asselbergs FW, van der Harst P, Linschoten M

Van Weel-Bethesda Hospital, Dirksland, the Netherlands: Wu KW

Zaans Medical Center, Zaandam, the Netherlands: Verschure DO

Zuyderland Medical Center, Heerlen, the Netherlands: Kietselaer BLJH

## Author contributions

Idea and study design: KRS, LAG; Data collection and analysis: JES, YZ, GX, WM, HS, K-LM, C-FC, L-XK, K-QW, JL, MA, KRS; Writing the article: JES, KRS; Draft revision: JES, GX, WM, HS, K-LM, C-FC, L-XK, K-QW, JL, MA, MR, LAG, KRS; All authors have read the manuscript and have approved this submission.

## Ethics approval and consent to participate

The meta-analysis portion of this project was reviewed by the Human Ethics Research Office, The University of Queensland (CRICOS provider number: 00025B; application number: 2020001352) and deemed to be exempt from ethics review under the National Statement on Ethical Conduct in Human Research (National Statement §5.1.22).

